# External validation of 4C ISARIC mortality score in the setting of a Saudi Arabian ICU. Retrospective study

**DOI:** 10.1101/2021.08.16.21262104

**Authors:** Shahzad A. Mumtaz, Saima A. Shahzad, Intekhab Ahmed, Mohammed A. Alodat, Mohamed Gharba, Zohdy A. Saif, Ahmed F. Mady, Waqas Mahmood, Huda Mhawish, Majd M. Abdulmowla, Waleed Aletreby

**Affiliations:** Critical Care Department, King Saud Medical City, Riyadh, Saudi Arabia; Critical Care Department, The Wollongong Hospital, NSW, Australia; Anesthesia Department, Faculty of Medicine, Tanta University, Tanta, Egypt; Faculty of Medicine, Alfaisal University, Riyadh, Saudi Arabia

**Keywords:** ISARIC, COVID-19, Mortality, Prediction, ICU

## Abstract

COVID-19 pandemic has burdened healthcare systems, necessitating the development of mortality prediction scores to guide clinical decisions and resource allocation. 4C ISARIC mortality score was developed and validated on a British cohort.

**Objectives:** External validation of the score in the setting of a large Saudi Arabian ICU.

**Method:** Retrospective chart review of COVID-19 patients admitted to ICU of King Saud Medical City, Riyadh, Saudi Arabia. Collecting data to calculate the score, then fitting a ROC curve against known patients’ outcome.

**Results:** Cohort included 1493 patients with 38% mortality, AUC of the score was 0.81 (95% CI: 0.79 – 0.83, p < 0.001), correctly classifying 72.67% of the cohort. Cut-off value of > 9 had sensitivity of 70.5% (95% CI: 66.6 – 74.3), specificity 73.97% (95% CI: 71 – 76.8), positive predictive value 62.4% (95% CI: 59.5 – 65.2), and negative predictive value 80.2% (95% CI: 78.2 – 82.4).

**Conclusion:** 4C ISARIC mortality risk score performed well with a good discriminatory ability for critically ill patients admitted to ICU in our setting. Cut-off > 9 was the optimal criterion.

## Introduction

Late 2019 the city of Wuhan (China) reported cases of unknown origin pneumonia that were identified to be due to infection with a novel coronavirus (1), the seventh member of the subfamily “Orthocoronavirinae” of enveloped RNA corona viruses, which is commonly named COVID-19 (2). As the transmission of COVID-19 was much higher than previous outbreaks due to other corona viruses (3), it quickly spread worldwide, to be declared a pandemic by the World Health Organization (WHO) on March 11, 2020 (4). As of August 6, 2021 the WHO reports more than 200 million confirmed cases, and more than four million deaths worldwide (5).

The spectrum of clinical presentation of COVID-19 infection is quite ample, with the majority of cases being asymptomatic, to mild cases requiring minimal support, up to critically ill cases requiring intensive care unit (ICU) admission, and mechanical ventilation (6). Such life-threatening COVID-19 presentations are commonly characterized by acute respiratory distress syndrome (ARDS), multi-organ failure, hyper-inflammatory responses, thrombo-embolic manifestations, among others (7). Accordingly, it is comprehendible that COVID-19 patients particularly the severe and critically ill ones have a high mortality rate (8), particularly in the presence of risk factors of poor prognosis identified in several studies such as comorbidities, elevated inflammatory biomarkers, and old age (3, 7).

In view of the variability of presenting pictures of COVID-19 and their progression, and the increasing burden the pandemic imposes on healthcare systems (9), the necessity arose for a tool of risk stratification, to allow early identification of COVID-19 patients at higher risk of mortality, using readily available objective criteria (8-10). A recent large size prospective cohort study (8) utilized The International Severe Acute Respiratory and emerging Infections Consortium (ISARIC) World Health Organization (WHO) Clinical Characterisation Protocol to develop and validate such a tool (ISARIC 4C Mortality Score, hereafter referred to as ISARIC) that seemed to outperform other risk stratification models (11). However; the tool’s development and validation on a United Kingdom only population, led the authors to question its generalizability, and invite validation of the score on settings outside the UK. Hence; this study was performed to validate the performance of the ISARIC score within the setting of a Saudi Arabian ICU.

## Method

This was a retrospective chart review observational study, carried out in the ICU at King Saud Medical City (KSMC), Riyadh, Saudi Arabia. KSMC is the largest government hospital in the central region of Saudi Arabia, it has a power of 1200 in-patient beds, with an ICU of 127 beds originally, however; since the beginning of the COVID-19 pandemic, and the designation of KSMC as a referral center for COVID-19 cases, the ICU’s capacity was increased to harbor 300 beds. The ICU is closed operated round the clock by intensivists, with a nurse: patient ratio of 1:1, almost half of the beds were converted to single rooms, and all beds are fully equipped with capabilities of invasive and non-invasive monitoring and ventilation. The ICU generally follows guidelines of COVID-19 management recommended by the Saudi Ministry of Health (12). The study was approved by the local institutional review board, with waiver of consent in view of its observational design.

### Patients and setting

This study included all patients admitted to our ICU between January 1, 2020 and June 30, 2021, as long as they were COVID-19 positive, confirmed by reverse transcription – polymerase chain reaction (RT-PCR), and of adult age (18 years or above). We excluded pregnant ladies, patients known to have pulmonary tuberculosis (PTB), and human immune-deficiency virus (HIV) positive patients. After the completion of data collection, we further excluded patients with missing variables that preclude the calculation of ISARIC score.

### Data management

The following variables were collected from the electronic database of our ICU and/or manually from the patients’ medical records: Age, gender, number of comorbidities, respiratory rate at hospital admission, peripheral oxygen saturation (SpO2) on room air at hospital admission, Glasgow Coma Scale (GCS) at hospital admission, first available blood urea level (mmol/L) and C-reactive protein (CRP) (mg/L). This data enabled us to calculate the ISARIC score based on the method of calculation described in the development and validation study (8) (Table S1). We additionally recorded ethnicity, smoking status, and nature of comorbidities.

### Outcomes

The primary outcome of the study was the performance of ISARIC score in our settings, by evaluating its discriminatory ability of all-cause hospital mortality against known outcomes. Secondary outcomes included hospital mortality, ICU LOS, and survival comparison between subjects with score above and below the optimal criterion derived from the analysis, in addition to sensitivity, specificity, positive predictive value (PPV) and negative predictive value (NPV) of the identified criterion.

### Statistical method

For descriptive purposes, we separately presented demographic and clinical characteristics for survivors and non-survivors, summarized as mean ± standard deviation (SD) for continuous variables, and frequency and percentage (%) for discrete variables. Continuous variables were compared by student t test or Wilcoxon rank sum test as appropriate, whereas, discrete variables were compared by chi square test or Fisher’s exact test as appropriate, comparison results were presented with corresponding 95% confidence interval (CI) and p value. The primary outcome was evaluated by fitting Receiver Operator Characteristic (ROC) curve using known outcomes as reference variable, and ISARIC score as classification variable, via 1000 bootstrap with replacement. We presented the area under the curve (AUC) and significance p value. Furthermore, the analysis identified the optimal criterion associated with Youden’s index, which was used as a cut-off value for the ICU survival analysis, presented as Kaplan Meier curve and Log-rank p value, in addition to sensitivity and specificity contingency table.

All tests were two tailed, and considered statistically significant if p value < 0.05. Statistical tests were performed by commercially available software STATA® [StataCorp. 2019. *Stata Statistical Software: Release 16*. College Station, TX: StataCorp LLC.].

## Results

During the study period there were 1749 ICU admissions of COVID-19 confirmed cases, we excluded 32 patients less than 18 years old, 12 pregnant ladies, 5 PTB, and 2 HIV positive cases. Furthermore, we excluded 205 (11.7%) patients missing CRP values precluding score calculation. Accordingly, the study enrolled 1493 patients with complete data and known hospital outcome.

Table 1 depicts the demographic and clinical characteristics of the cohort, and comparison between survivors and non-survivors. There were 567 (38%) hospital deaths in the cohort, different mortality rates across categories of ISARIC score are shown in Figure S1. The cohort had a mean age of 53.2 ± 14.1 years, with a majority (76.3%) of males. Survivors had significantly different values than non-survivors in all variables used in the calculation of ISARIC score, in the direction of a lower score, with the exception of gender distribution, and significantly lower prevalence of all recorded comorbidities except for chronic kidney disease, ischemic heart disease, and smoking. Majority of the patients were South-East Asians (42%) followed by Saudis (28.2%) then Arab Non-Saudi patients (24.4%). Only Saudis and South-East Asians had significantly higher survival than mortality rates (Figure S2). The average ICU LOS of the cohort was 10.5 ± 9 days, being significantly shorter for survivors compared to non-survivors (8.1 ± 7.2 VS 14.3 ± 10.2, 95% CI of difference: -7 to -5.3, p < 0.001) (Figure S3). The cohort had a mean ISARIC score of 8.6 ± 5.3, survivors had significantly lower score than non-survivors (6.3 ± 4.3 Vs 12.4 ± 4.3, 95% CI of difference: -6.5 to -5.6; p < 0.001).

**Table 1:** Demographic and clinical characteristics of the cohort, and comparison between survivors and non-survivors:

| Variable | All cohort<br>(n = 1493) | Survivors<br>(n = 926) | Non-Survivors<br>(n = 567) | 95% CI of<br>difference | P value |
| --- | --- | --- | --- | --- | --- |
| Age (mean ± SD) | 53.2±14.1 | 51.1±13.6 | 56.8±14.4 | -7.2 to -4.3 | <0.001 |
| Males: n (%) | 1139<br>(76.3%) | 708 (76.5%) | 431<br>(76%) | -4% to 5.1% | 0.8 |
| Ethnicity: n (%) |  |  |  |  |  |
| Saudi | 421 (28.2%) | 295 (31.9%) | 126 (22.2%) | 5% to 14.3% | <0.001 |
| Arab Non-Saudi | 365 (24.4%) | 241 (26%) | 124 (21.9%) | -0.5% to 8.6% | 0.08 |
| South-east Asian | 627 (42%) | 338 (36.5%) | 289 (51%) | 9.2% to 19.7% | < 0.001 |
| African | 45 (3%) | 33 (3.6%) | 12 (2.1%) | -0.4% to 3.2% | 0.1 |
| Other | 6 (0.4%) | 4 (0.4%) | 2 (0.4%) | -0.7% to 1% | 0.7 |
| Unknown | 29 (2%) | 15 (1.6%) | 14 (2.4%) | -0.7% to 2.6% | 0.4 |
| Number of comorbidities<br>(mean ± SD) | 1.5 ± 1.1 | 1.3 ± 1.1 | 1.7 ± 1.2 | -0.5 to -0.2 | < 0.001 |
| Respiratory Rate (mean ± SD) | 28.3±9.6 | 22.4±6.8 | 37.8±4.6 | -16 to -14.8 | < 0.001 |
| Peripheral oxygen saturation<br>(mean ± SD) | 92.4 ± 3.4 | 92.9 ± 3.3 | 91.6 ± 3.4 | 0.96 to 1.7 | < 0.001 |
| Glasgow Coma Scale (mean ± SD) | 14.6 ± 0.9 | 14.7 ± 0.8 | 14.5 ± 0.9 | 0.1 to 0.3 | < 0.001 |
| Blood Urea (mean ± SD) | 9.8 ± 5.4 | 6.7 ± 2.9 | 14.7 ± 4.8 | -8.3 to -7.6 | < 0.001 |
| C-Reactive Protein (mean ± SD) | 75.2 ± 33.2 | 51.8 ± 17.1 | 112 ± 12.9 | -61.8 to -58.6 | < 0.001 |
| Smoker: n (%) | 766 (51.3%) | 458 (49.5%) | 308 (54.3%) | -0.5% to 10.1% | 0.08 |
| Diabetes Mellitus: n (%) | 815 (54.6%) | 458 (49.5%) | 357<br>(63%) | 8.2% to 18.7% | <0.001 |
| Hypertensive: n (%) | 780 (52.2%) | 443 (47.8%) | 337<br>(59.4%) | 6.3% to 16.8% | < 0.001 |
| Asthma/COPD: n(%) | 74<br>(5%) | 37<br>(4%) | 37<br>(6.5%) | 0.1% to 5.1% | 0.04 |
| Chronic Kidney Disease: n(%) | 235 (15.7%) | 139 (15%) | 96 (16.9%) | -2% to 5.9% | 0.4 |
| Ischemic Heart Disease: n(%) | 277 (18.6%) | 154 (16.6%) | 123 (21.7%) | -0.8% to 11.6% | 0.09 |
| ICU LOS (mean ± SD) | 10.5 ± 9 | 8.1 ± 7.2 | 14.3 ± 10.2 | -7 to -5.3 | < 0.001 |
| ISARIC Score<br>(mean ± SD) | 8.6 ± 5.2 | 6.3± 4.3 | 12.4± 4.3 | -6.5 to -5.6 | < 0.001 |

COPD = chronic obstructive pulmonary disease, ICU = intensive care unit, LOS = length of stay, SD = standard deviation, CI = confidence interval

### Primary outcome

The fitted ROC curve of ISARIC score against known outcomes had AUC of 0.81 (95% CI: 0.79 – 0.83, p < 0.001) (Figure 1). The associated criterion with Youden’s index was a score more than 9, accordingly, survival based on that cut-off value estimated by Kaplan Meier curve showed a median survival of 22 days with ISARIC score ≤ 9, while a median survival of 16 days with scores > 9, the associated Log-Rank test was highly significant (Figure 2). Sensitivity associated with score >9 was 70.5% (95% CI: 66.6 – 74.3), speceficity was 73.97% (95% CI: 71 – 76.8), PPV of 62.4% (95% CI: 59.5 – 65.2), and NPV of 80.4% (95% CI: 78.2 – 82.4), and the cut-off value correctly classified 72.7% of the cohort. (Table S2).

**Figure 1:**
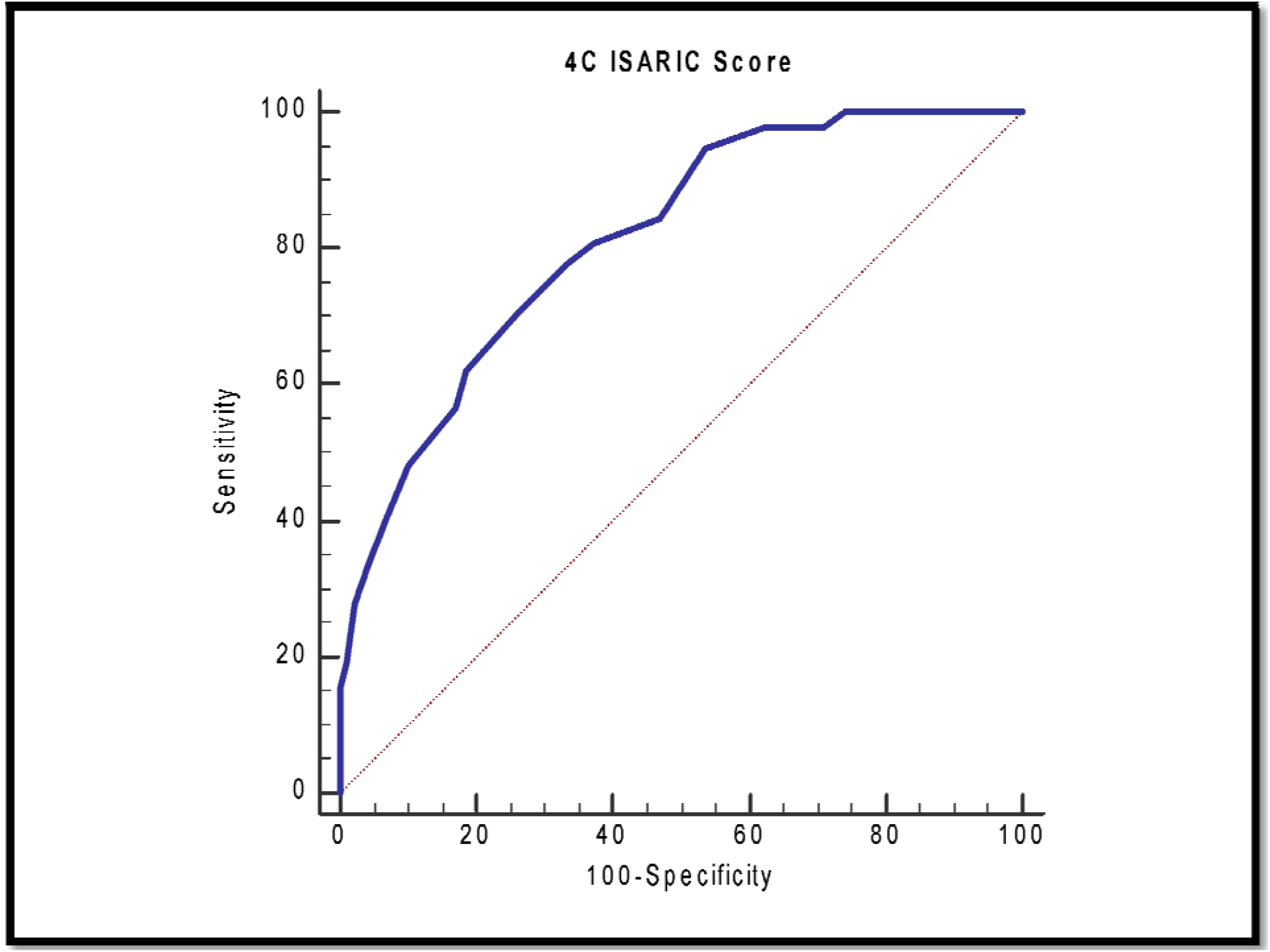
Area under the curve of 4C ISARIC Score discriminatory ability: AUC = 0.81 (95% CI: 0.79 – 0.83, p < 0.001)

**Figure 2:**
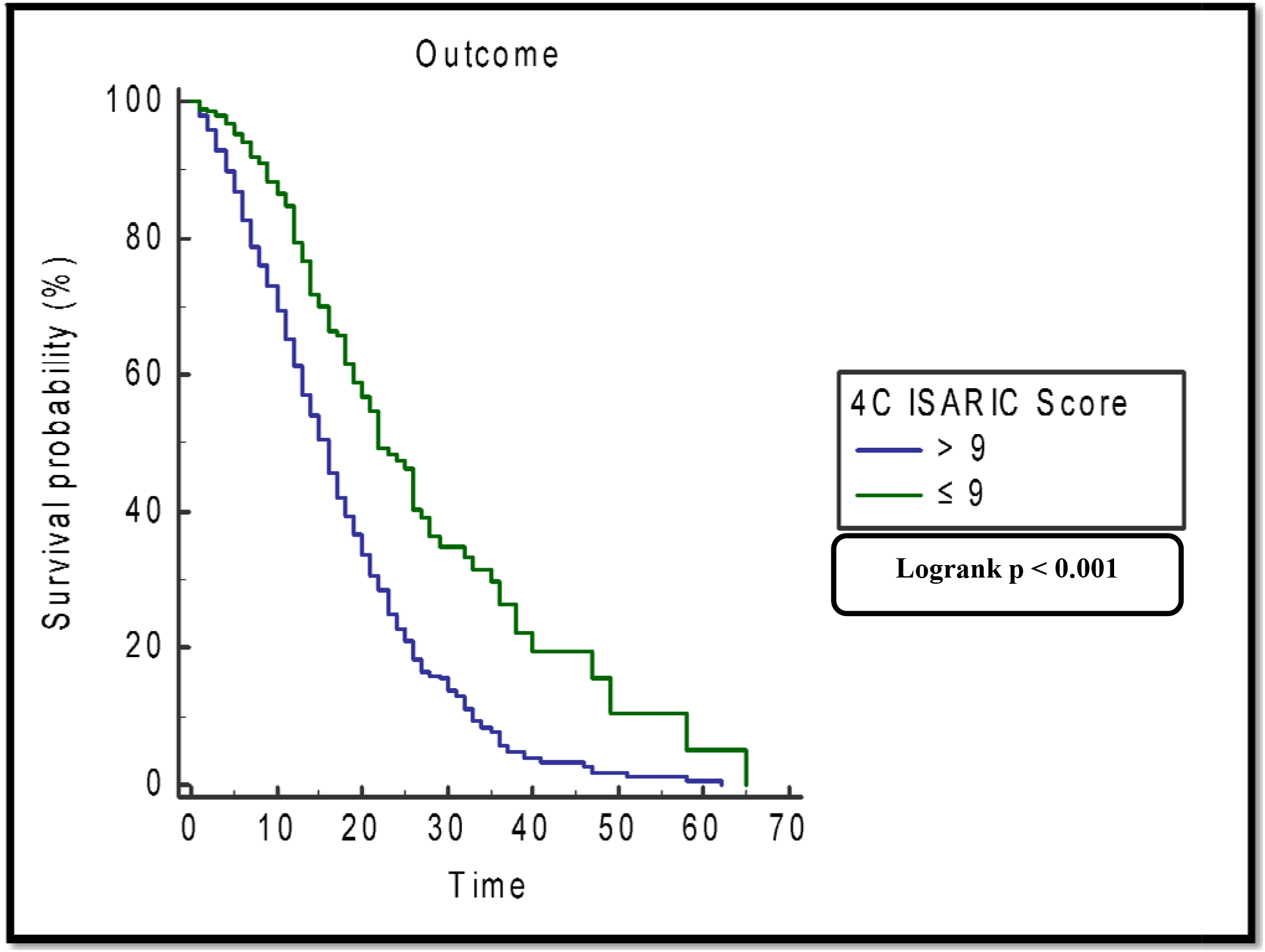
Kaplan Meier Survival based on Cutoff of > 9:

### Secondary outcomes

Dividing the enrolled patients into two groups based on ISARIC score of ≤ 9 and > 9 showed significantly higher mortality in the > 9 group compared to ≤ 9 (62.4% Vs 19.6%, 95% CI: 38% to 47.4%; p < 0.001), similarly group ≤ 9 had ICU LOS of 9.1 ± 8.2 days, significantly shorter than that of group > 9 with LOS of 12.3 ± 9.7 days (95% CI: 2.3 to 4.1, p < 0.001). Table S3.

## Discussion

Since the beginning of the COVID-19 pandemic healthcare systems have been overwhelmed with increasing numbers of patients on one side, and limited resources on the other (3), particularly, with proportions of critically ill patients requiring mechanical ventilation and/or ICU admission in the vicinity of 15% (13) up to more than 30% in some reports (14). Hence, the need arose for a prediction model that can predict outcomes of COVID-19 patients as early and as accurately as possible, in order to aid clinical decision making about aggressive treatment and resource allocation (8). Such a score was developed and validated on a large cohort, utilizing eight readily available variables upon hospital admission, namely: age, gender, number of comorbidities, peripheral oxygen saturation, respiratory rate, level of consciousness, C-reactive protein and urea levels (8). The so called 4C ISARIC Mortality Score is not only easy to calculate, but it also combines variables reflecting the patients’ demography, comorbidity, physiological current condition, and lab investigations, while avoiding parameters that rely on radiological imaging or only become available after ICU admission (15, 16). The model showed a high discriminatory ability for in-hospital mortality (AUC = 0.786; 95% CI: 0.781 – 0.79), and the authors were able to categorize patients into four categories of severity with a uniformly increasing mortality risk (Table S4). However; a limitation of the model was that the development and validation cohorts were entirely of the United Kingdom population, which led the authors to invite external validation on populations outside the UK.

We carried out our study on a population just shy of 1500 patients, since we chose to exclude cases with missing data (11.2%) precluding calculation of ISARIC score, although modern imputation methods (such as multiple imputation) are likely to produce valid estimates, but only when critical assumptions have been met (17) which may have not been true for our data, they still carry the potential to cause bias (18). Enrolled patients in our analysis were of multiple ethnicities, representing the general population of Saudi Arabia, accordingly, it provided a good external validation cohort for the ISARIC score.

In our study ISARIC score had an excellent (19) discrimination ability for mortality risk, AUC of 0.81 (95% CI: 0.79 – 0.83, p < 0.001), this was slightly higher than that reported by the original development and validation study, although uncommon in external validation studies, this seems to be mirrored by others, PMEL et al (9) and Wellbelove et al (20) reported an even higher AUC of 0.84 (95% CI: 0.79 – 0.88) and 0.83 (95% CI: 0.71 – 0.95) respectively, the latter with a wider CI possibly due to smaller sample size. Other reports presented AUC values much closer to the validation study (21), and lower (22). This variation among studies – although minimal – utilizing the same prediction model may be a reflection of the variations in the studied populations themselves, with regards to their demographic characteristics, clinical severity, and even sample size. For example, the population in our study included several ethnic groups, a wide range of age (lowest 18 and highest 104 years), unequal gender distribution with majority of males (76.3%), furthermore, our cohort included only patients admitted to the ICU, which means they presented a more critical picture of the disease, while the original study enrolled all hospital admissions, some of whom may have not been as critical. Regardless of those variations, all reports including our study definitely place the discriminatory ability of ISARIC score within the area of acceptable to excellent.

Our external validation – similar to the original study – showed rising mortality rates across groups of severity (Figure S1, Table S4) indicating good performance of the model by showing higher mortality risk as the score increases, despite having higher mortality rates within each group, once more probably reflecting the criticality and severity of our cohort. In our analysis, the optimal cut-off value associated with Youden’s index was a score > 9, this value correctly classifies 72.7% of the cohort. The diagnostic parameters (sensitivity, specificity, PPV, and NPV) of the cut-off of >9 in our model were considerably lower compared to the same cut-off value in the British study, perhaps as a result of inclusion of only critically ill patients, which makes the prediction of their outcome much more difficult than mild cases, since critically ill COVID-19 patients are subject to many complications (7) that could be the indirect cause of death, this is easy to comprehend when we compare the higher mortality rate (38%) in our study to the lower rate of (32.2%) in the original study. Possibly one of the most important diagnostic parameters is the NPV, which translates into the probability of not dying if the patient does not have a score of >9, in our model NPV was 80.2% (95% CI: 78.2 – 82.4), which provides a reasonable risk probability to guide clinical decision making, this discrimination was supported by our finding of a significantly higher survival of patients with a score ≤ 9, shorter ICU LOS, and lower mortality.

### Limitations

Our study suffers several limitations, first, the inherent limitation within the retrospective design, second, this remains a single center study, reflecting the management in only our ICU, third, our choice not to use imputation methods on missing data may have reduced our sample size and impacted our results, fourth, despite working with a quite diverse population, our cohort lacked several ethnicities, so our results may not be applicable in other regions of the world, and finally, generalizability of our results may be limited to only critically ill patients, as we didn’t include non-ICU admissions.

## Conclusion

4C ISARIC Mortality score is a quick and easy to use mortality risk prediction model of COVID-19 patients, with an acceptable performance for critically ill patients admitted to ICU in our setting. A cut-off value of > 9 has the best negative predictive value.

## Data Availability

Data available with corresponding author upon justified request.

## Acknowledgment

We would like to express gratitude to:

Ms. Katrina Baguisa.

Ms. Rehab Alfenaikh

Ms. Ika Fabriantini

