## Supplementary file for "External validation of 4C ISARIC mortality score in the setting of a Saudi Arabian ICU. Retrospective study"

**Table S1: Calculation of 4C ISARIC Mortality Score.**

**Table S2: Contengency table of Youden’s index associated criterion.**

**Table S3: Comparison of patients by cut-off value of ISARIC score > 9.**

**Table S4: Categories and risk of mortality of ISARIC Score, in the original and current study.**

**Figure S1: Distribution of mortality rate by categories of 4C ISARIC Score Severity.**

**Figure S2: Outcome distribution by Ethnicity.**

**Figure S3: ICU LOS distribution, by Outcome.**

Table S1: Calculation of ISARIC 4C Mortality Score:

| Variable | Score |
| --- | --- |
| **Age Group:**  Less than 50  50 – 59  60 – 69  70 – 79  80 or more | 0  2  4  6  7 |
| **Gender:**  Male  Female | 1  0 |
| **Number of comorbidities:**  0  1  2 or more | 0  1  2 |
| **Respiratory Rate (breath / min):**  Less than 20  20 – 29  30 or more | 0  1  2 |
| **Peripheral oxygen saturation on room air:**  Equal to or more than 92%  Less than 92% | 0  2 |
| **Glasgow Coma Scale:**  15  Less than 15 | 0  2 |
| **Blood Urea (mmol/L):**  Less than 7  7 – 14  More than 14 | 0  1  3 |
| **C – Reactive Protein (mg/L):**  Less than 50  50 – 99  100 or more | 0  1  2 |

Data are recorded upon hospital admission

Table S2: Contengency table of Youden’s index associated criterion:

|  |  | **Actual** | |  |
| --- | --- | --- | --- | --- |
|  |  | **Dead** | **Alive** | **SUM** |
| **ISARIC Score>9** | **Dead** | **400** | **241** | **641** |
|  | **Alive** | **167** | **685** | **852** |
|  | **SUM** | **567** | **926** | **1493** |

Sensitivity = 70.5% (95% CI: 66.6 – 74.3)

Speceficity = 73.97% (95% CI: 71 – 76.8)

PPV = 62.4% (95% CI: 59.5 – 65.2)

NPV = 80.2% (95% CI: 78.2 – 82.4)

Correctly classified: 72.67%

Table S3: Comparison of patients by cut-off value of ISARIC score > 9:

| Variable | Score ≤ 9 (n=852) | Score > 9  (n = 641) | 95% CI of difference | P value |
| --- | --- | --- | --- | --- |
| Age (mean ± SD) | 46.8 ± 11.6 | 61.8 ± 12.4 | 60.8 to 62.7 | < 0.001 |
| Males: n (%) | 628 (73.7%) | 511 (79.7%) | 1.6% to 10.3% | 0.008 |
| Saudis: n (%) | 421 (49.4%) | 10 (1.6%) | 44.2% to 51.3% | < 0.001 |
| DM: n (%) | 328 (38.5%) | 487 (76%) | 32.7% to 42.1% | < 0.001 |
| HTN: n (%) | 269 (31.6%) | 511 (80%) | 43.8% to 52.7% | < 0.001 |
| CKD: n (%) | 51 (6%) | 184 (28.7%) | 18.8% to 26.7% | < 0.001 |
| Asthma/COPD: n (%) | 33 (3.9%) | 41 (6.4%) | 0.2% to 5% | 0.04 |
| IHD: n (%) | 71 (8.3%) | 206 (32.1%) | 19.7% to 28% | < 0.001 |
| Smoking: n (%) | 422 (49.5%) | 344 (53.7%) | -1.02% to 9.4% | 0.1 |
| ICU LOS  (mean ± SD) | 9.1 ± 8.2 | 12.3 ± 9.7 | 2.3 to 4.1 | < 0.001 |
| Hospital Mortality n (%) | 167 (19.6%) | 400 (62.4%) | 38% to 47.4% | < 0.001 |

DM = diabetes mellitus, HTN = hypertension, CKD = chronic kidney disease, COPD = chronic obstructive pulmonary disease, IHD = ischemic heart disease, ICU = intensive care unit, LOS = length of stay, SD = standard deviation.

Table S4: Categories and risk of mortality of ISARIC Score, in the original and current study:

| 4C Mortality Score | Risk Group | Risk of In-hospital mortality | |
| --- | --- | --- | --- |
|  |  | Original Study* | Current Study |
| 0 - 3 | Low | 1.2 – 1.7 % | 2.3 – 7.3 % |
| 4 - 8 | Intermediate | 9.1 – 9.9 % | 20.8 – 28.9 % |
| 9 - 14 | High | 31.4 – 34.9 % | 43.9 – 52.7 % |
| ≥ 15 | Very high | 61.5 – 66.2 % | 76.6 – 86.8 % |

*Knight SR, Ho A, Pius R, Buchan I, Carson G, Drake TM, et al; ISARIC4C investigators. Risk stratification of patients admitted to hospital with covid-19 using the ISARIC WHO Clinical Characterisation Protocol: development and validation of the 4C Mortality Score. BMJ. 2020 Sep 9;370:m3339.

Figure S1: Distribution of mortality rate by categories of 4C ISARIC Score Severity:

|  | Count | MR |
| --- | --- | --- |
| Low (0-3) | 282 | 4.3 |
| Intermediate (4-8) | 462 | 24.7 |
| High (9-14) | 516 | 48.3 |
| Very High (≥15) | 233 | 82.1 |

Figure S2: Outcome distribution by Ethnicity:

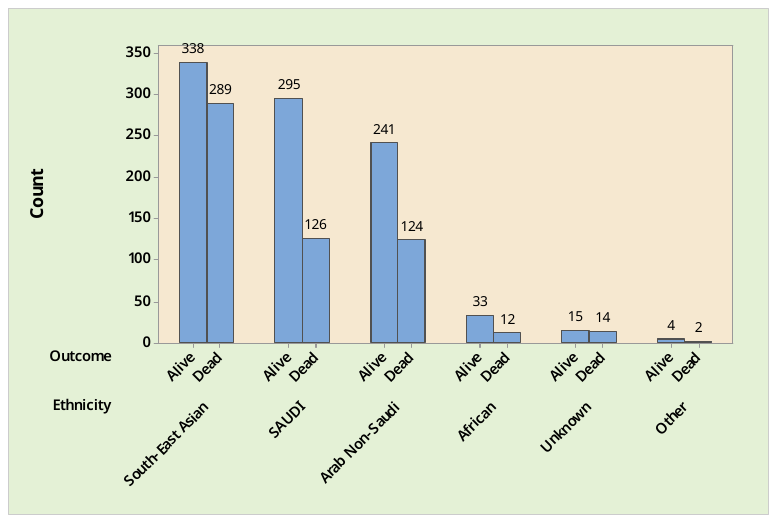

Figure S3: ICU LOS distribution, by Outcome:

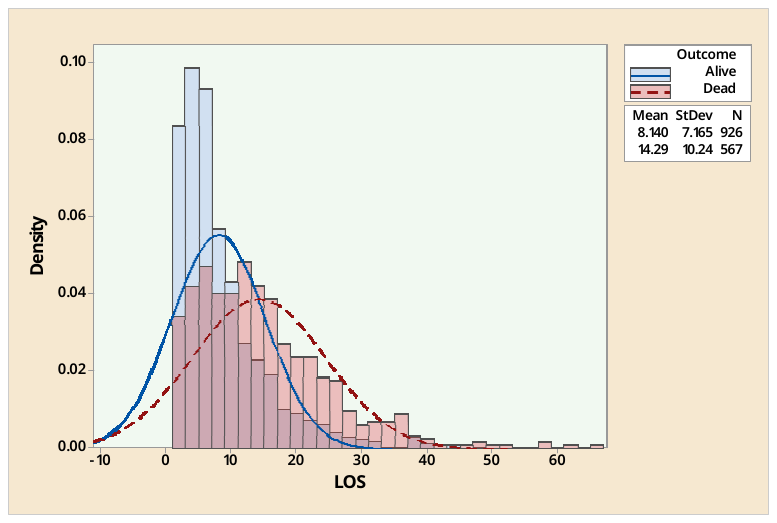
